# The Potential of Dietary Strategies: The Impact of Low-Carbohydrate Diet on Cardiovascular-Kidney-Metabolic Syndrome

**DOI:** 10.1101/2025.01.03.25319951

**Authors:** Zhuoxing Li, Xiunan Liu, Xin Ma, Mao Xiao, Xue Zhang, Yanyi Deng, Hao Liu, Yun Sun, Xiang Xiao

## Abstract

This study aims to explore the prevalence of Low-carbohydrate diet (LCD) among individuals with cardiovascular-kidney-metabolic (CKM) syndrome and its relationship with prognosis.

**Methods:** The individuals diagnosed with CKM were included from the National Health and Nutrition Examination Survey (NHANES) database between 2009 and 2018. Participants were divided into two groups: those on a LCD and those not on a LCD. Multivariate logistic regression analysis was conducted to assess the factors influencing the choice of LCD among CKM individuals. Kaplan-Meier survival curve analysis and multivariate Cox regression analysis were performed to evaluate the association between LCD and the risk of all-cause mortality. Stratified analysis was performed to assess the consistency of results.

**Results:** A total of 1000 non-CKM (stage 0) individuals and 11,141 CKM individuals (stages 1, 2, 3, and 4) were included. The proportions of individuals on LCD for CKM stages 0, 1, 2, 3, and 4 were 1.16%, 2.49%, 1.94%, 1.24%, and 1.68%, respectively. Multivariate logistic regression analysis indicated that age, and race might influence the choice of LCD among CKM individuals. Multivariate Cox regression analysis revealed that compared to non-LCD individuals, those on LCD had a 62% reduced risk of all-cause mortality (HR = 0.38; 95% CI, 0.15-0.92, P = 0.03). Stratified analysis showed no interaction between LCD and CKM stages (*P* for interaction < 0.05).

**Conclusion:** The proportion of individuals on LCD in CKM is relatively low. Furthermore, LCD can decrease the risk of all-cause mortality among CKM individuals, regardless of CKM stage.

## Introduction

Cardiovascular-kidney-metabolic (CKM) Syndrome is a series of systemic diseases caused by the pathophysiological interactions between metabolic abnormalities, chronic kidney disease (CKD), and cardiovascular disease (CVD), proposed by the American Heart Association (AHA) in 2023 ^1^.In recent years, with changes in lifestyle, the incidence of CKM has been continuously increasing; by 2020, nearly 90% of American adults met the criteria for CKM syndrome, making it a significant public health issue ^2^. CKM not only increases the risk of CVD, chronic kidney disease, and metabolic diseases (such as type 2 diabetes), but also significantly raises the risk of mortality for individuals ^3^.

Dietary interventions are an important component in the treatment of metabolic diseases ^4^. The AHA identifies several influencing factors of CKM including social determinants of health (SDOH), one of which is dietary intake. Inappropriate dietary intake can further affect the likelihood of CKM syndrome and lead to adverse outcomes ^5^. LCD is recognized as a healthy dietary pattern by the ADA, American Heart Association, and Canadian Diabetes Association ^6^. This dietary approach has been utilized since the last century to reduce the frequency of seizures and has been validated and applied in the management of diseases across cardiovascular, neurological, and endocrine systems ^7^. LCD (including very low carbohydrate or ketogenic diets) is defined as a dietary pattern where the total carbohydrate intake is less than 26% of total daily caloric intake ^8^. However, LCD requires the involvement of a multidisciplinary team (such as physicians, nurses, and nutritionists), and adverse reactions, dietary restrictions, and unpleasant taste may lower individuals’ motivation and lead to treatment discontinuation ^9^. Therefore, the ability to adhere to a long-term LCD poses a significant challenge. There are currently no reported studies assessing the proportion and adherence to LCD among individuals with CKM.

Furthermore, research results regarding the impact of LCD on the prognosis of cardiovascular, renal, and metabolic syndromes are inconsistent, with some studies yielding contrary conclusions. Although cardiovascular and endocrine risk factors have been improved in individuals on LCD ^10, 11^, most studies have not observed a significant correlation with total CVD, all-cause mortality, or cardiovascular mortality ^12^. Concurrently, evidence suggests that LCD offer little benefit in controlling CKD and its complications ^13^; however, LCD scores among CKD individuals exhibit a significant positive correlation with all-cause mortality ^14^. Moreover, multiple studies have demonstrated that LCD can reduce all-cause mortality risk in individuals with obesity, diabetes, and other metabolic diseases ^15^. Nonetheless, there is a scarcity of research exploring the relationship between LCD and all-cause mortality risk in individuals with CKM.

Thus, this study aims to leverage data from the NHANES database to understand the current status of LCD among individuals with CKM, potential influencing factors of LCD adherence, and the relationship between these dietary patterns and individual prognosis, providing important references for guiding personalized dietary management strategies for CKM individuals.

## 1 Methods

### 1.1 Study Design and Participants

This study included individuals from the National Health and Nutrition Examination Survey (NHANES) database collected between 2009 and 2018. The core objective of the NHANES database is to conduct in-depth interviews and standardized physical examinations conducted by Mobile Examination Centers (MEC) to comprehensively assess the health and nutritional levels of the non-institutionalized U.S. population reflecting individual demographic characteristics, health status, and nutritional data ^16^.

After filtering, individuals meeting the AHA-defined stages of CKM (stages 0-4) were included. We adopted the staging structure proposed by the AHA to ensure the accuracy and reliability of the results: 1) Stage 0, no CKM risk factors; 2) Stage 1, excessive obesity, dysfunctional obesity, or the presence of both; 3) Stage 2, metabolic risk factors, moderate to high-risk CKD, or the presence of both; 4) Stage 3, subclinical CVD overlapping with CKM risk factors, very high-risk CKD, or high predicted CVD risk; 5) Stage 4, clinical CVD coexisting with CKM risk factors ^1^ **(Supplementary Table 1)**. Finally, non-CKM was defined as stage 0 and CKM as stages 1-4. LCD was defined as an energy intake from carbohydrates less than 26% of total energy or a daily carbohydrate intake of less than 130g ^17^ **(Supplementary Table 1)**. Exclusion criteria included: 1) age <20 years, 2) pregnancy, 3) missing data related to LCD, and 4) missing CKM diagnosis **(Figure 1)**. All-cause mortality refers to the termination of life due to any cause ^18^. Definitions regarding smoking, alcohol consumption, hypertension, anemia, hyperlipidemia, diabetes, chronic kidney disease, and CVD are detailed in **Supplementary Table 1**. eGFR was measured using the Chronic Kidney Disease Epidemiology Collaboration (CKD-EPI) equation^19^.

The NHANES project is a survey implemented by the National Center for Health Statistics (NCHS) of the Centers for Disease Control and Prevention (CDC) and has received formal authorization from the Institutional Review Board of the NCHS. This study adheres to the ethical guidelines established by the Declaration of Helsinki of 1964 and its subsequent amendments, and has undergone rigorous approval by the National Center for Health Statistics Research Ethics Review Board. All participants in the survey signed informed consent documents based on a comprehensive understanding of the study ^20^.

### 1.2 Statistical Analysis

Following the guidelines of the CDC, this study adequately considered sample weights in the statistical analysis, using means and their standard errors (SE) to describe continuous variables and percentages along with their SE to describe categorical variables. We utilized a complex sampling design logistic regression model to explore the factors influencing individual choice of LCD; employed the complex sampling Kaplan-Meier survival analysis to evaluate the association between LCD and overall mortality prognosis in individuals with CKM; and used a complex sampling multivariable Cox regression model to investigate the relationship between LCD and the risk of all-cause mortality, as well as cardiovascular mortality in individuals with CKM. Furthermore, we performed stratified analyses to verify the robustness of the results. For missing values, we employed multiple imputation methods and excluded samples with missing data for sensitivity analysis. To avoid biases that might arise from eating issues due to certain diseases, we excluded the following factors in the sensitivity analysis: gastrointestinal malignancies (such as esophageal cancer, gallbladder cancer, rectal cancer, and liver cancer). All statistical analyses were conducted using R software version 4.3.1, with a two-sided P value of less than 0.05 considered the threshold for statistical significance.

## 2 Results

### 2.1 Baseline Characteristics

Our study included data from 49,693 individuals from the NHANES database collected between 2009 and 2018. After exclusions, a total of 1000 individuals Non-CKM (stage 0) and 11,141 individuals with CKM (stages 1, 2, 3, and 4) were included in the final analysis. Among CKM individuals, the proportion of stage 1 individuals was 24.66%, stage 2 individuals were 60%, stage 3 individuals were 4.24%, and stage 4 individuals was 11.1%. The average age was 50.04 years, with males constituting 49.76%. The mean body mass index (BMI) was 29.98kg/m^2^, and the proportions of smokers and drinkers were 46.72% and 88.77%, respectively. The prevalence rates of hyperlipidemia, anemia, hypertension, diabetes, and CKM were 73.56%, 7.17%, 47.64%, 18.87% and 15.650%, respectively **(Table 1)**.

### 2.2 Status of LCD among CKM Individuals

Among the 11,141 CKM individuals included, 1.71% (191/11141) were classified as LCD. Compared to non-LCD individuals, LCD individuals had higher income (*P* < 0.05) **(Figure 1, Table 1).** The proportion of LCD individuals under 65 years old was 2.16%, while for those aged 65 years and above, it was 1.53%, with no significant difference between the two groups (*P* = 0.13); the proportion of LCD individuals among males was 1.83% and among females was 2.21% (*P* = 0.38), indicating no significant difference between the two groups. The proportions of LCD individuals from Mexican American, Non-Hispanic Black, Non-Hispanic White, Other Hispanic, and Other Race-Including Multi-Racial categories were 0.7%, 2.43%, 2.15%, 2.27%, and 1.66%, respectively, with no significant differences observed between groups (*P* = 0.16). The proportions of individuals in CKM stages 0, 1, 2, 3, and 4 were 1.16%, 2.49%, 1.94%, 1.24%, and 1.68%, respectively (*P* = 0.54) **(Table 2).**

### 2.3 Factors Influencing the Choice of LCD

After adjusting for factors such as age (‘<65’ years, ‘≥65’ years), gender (‘Female’, ‘Male’), race (‘Mexican American’, ‘Non-Hispanic Black’, ‘Non-Hispanic White’, ‘Other Hispanic’, ‘Other Race - Including Multi-Racial’), BMI (<30kg/m^2^, ≥30kg/m^2^), smoking status (‘No’ or ‘Yes’), alcohol use (‘No’ or ‘Yes’), education level (‘College or above’, ‘High school or equivalent’, ‘Less than high school’), poverty status (‘0-1’, ‘1.1-3’, ‘>3’), anemia (‘No’ or ‘Yes’), CKD (‘No’ or ‘Yes’), hyperlipidemia (‘No’ or ‘Yes’), hypertension (‘No’ or ‘Yes’), diabetes mellitus (DM) (‘No’ or ‘Yes’), and cardiovascular disease (CVD) (‘No’ or ‘Yes’), both age and race could influence the choice of CKM individuals regarding LCD **(All *P* < 0.05) (Supplementary-table 2)**.

### 2.4 Risk of All-Cause Mortality

#### 2.4.1 KM Survival Analysis

Kaplan-Meier survival analysis was conducted to assess the relationship between LCD and the risk of all-cause mortality in CKM individuals. After adjusting for age (‘<65’ years, ‘≥65’ years), gender (‘Female’, ‘Male’), race (‘Mexican American’, ‘Non-Hispanic Black’, ‘Non-Hispanic White’, ‘Other Hispanic’, ‘Other Race - Including Multi-Racial’), BMI (<30 kg/m^2^, ≥30 kg/m^2^), smoking status (‘No’ or ‘Yes’), alcohol use (‘No’ or ‘Yes’), education level (‘College or above’, ‘High school or equivalent’, ‘Less than high school’), poverty status (‘0-1’, ‘1.1-3’, ‘>3’), anemia (‘No’ or ‘Yes’), CKD (‘No’ or ‘Yes’), hyperlipidemia (‘No’ or ‘Yes’), hypertension (‘No’ or ‘Yes’), DM (‘No’ or ‘Yes’), and CVD (‘No’ or ‘Yes’), the results indicated that CKM individuals in the LCD group had better survival outcomes compared to those in the non-LCD group (Log-rank *P* < 0.001) **(Figure 2)**.

#### 2.4.2 Multivariate Cox Regression Analysis

After multivariate adjustment for age (‘< 65’ years, ‘≥ 65’ years), gender (‘Female’, ‘Male’), race (‘Mexican American’, ‘Non-Hispanic Black’, ‘Non-Hispanic White’, ‘Other Hispanic’, ‘Other Race - Including Multi-Racial’), BMI (< 30 kg/m^2^, ≥30 kg/m^2^), smoking status (‘No’ or ‘Yes’), alcohol use (‘No’ or ‘Yes’), education level (‘College or above’, ‘High school or equivalent’, ‘Less than high school’), poverty status(‘0-1’, ‘1.1-3’, ‘>3’), anemia (‘No’ or ‘Yes’), hyperlipidemia (‘No’ or ‘Yes’), hypertension (‘No’ or ‘Yes’), DM (‘No’ or ‘Yes’), CKD (‘No’ or ‘Yes’) and CVD (‘No’ or ‘Yes’), the results of the Cox regression analysis showed that the risk of all-cause mortality in CKM individuals in the LCD group was reduced by 62% compared to the non-LCD group (HR=0.38; 95% CI, 0.15-0.92, *P* = 0.03) **(Supplementary-table 3, 4; Figure 3)**.

#### 2.4.3 Stratified Analysis

In the stratified analysis, the relationship between LCD and the overall mortality risk of CKM individuals was assessed for potential influences from factors such as age (‘< 65’ years, ‘≥ 65’ years), gender (‘Female’, ‘Male’), BMI (< 30 kg/m^2^, ≥ 30 kg/m^2^), race (‘Mexican American’, ‘Non-Hispanic Black’, ‘Non-Hispanic White’, ‘Other Hispanic’, ‘Other Race - Including Multi-Racial’), DM (‘No’ or ‘Yes’), CKD (‘No’ or ‘Yes’), CVD (‘yes’ or ‘no’), and CKM stages (Stage 1-2, 3, and 4). The results indicated that, except for the DM (‘No’ or ‘Yes’) (*P* for interaction < 0.05), there were no interactions between LCD and variables such as age, gender, BMI, race, presence of CKD or CVD, and CKM stages (*P* for interaction > 0.05) **(Supplementary - table 5**, Figure 4).

#### 2.4.4 Sensitivity Analysis

To reduce the impact of missing values on the results, we performed data deletion and re-evaluated the relationship between LCD and the all-cause mortality risk among CKM individuals. After multivariable adjustment, the results of Cox regression analysis indicated that compared to the non-LCD group, the all-cause mortality risk in the LCD group was reduced by 67% (HR = 0.33; 95% CI, 0.13-0.85, *P* = 0.02) **(Supplementary table 6)**.

To mitigate the result bias caused by individual dietary factors, we excluded individuals with gastrointestinal malignancies. The results of multivariate Cox regression analysis showed that compared to the non-LCD group, the all-cause mortality risk in the LCD group was reduced by 62% (HR = 0.38; 95% CI, 0.15-0.91, *P* = 0.03) **(Supplementary table 7)**.

## 3 Discussion

This study found that the proportion of CKM individuals currently selecting LCD was very low. Factors such as age and race might affect CKM individuals’ choice of LCD. Additionally, LCD was associated with a reduced risk of all-cause mortality among CKM individuals, regardless of the stage of CKM.

Obesity is central to CKM syndrome. The AHA recommends that individuals with CKM stages 0-4 should intentionally reduce their weight through lifestyle and dietary changes, preferably through sustained interventions lasting ≥ 6 months ^5^. The American Diabetes Association (ADA) and the European Association for the Study of Diabetes (EASD) have certified that LCD has significant effects on reducing body weight and body fat percentage in patients with obesity or metabolic syndrome, particularly in terms of reducing abdominal and visceral fat ^21^. Therefore, the application of LCD among CKM individuals and its impact on prognosis are issues that need to be explored.

In recent years, LCD have garnered considerable attention and application across multiple disease domains. Among diabetic patients, the proportion of those adopting LCD has reached 20-40% ^22^. However, the percentage of individuals with CKM opting for LCDs was only 1.71%, a phenomenon that warrants greater recognition and attention. Meanwhile, there were no significant differences in the proportion of LCD adopted by patients of different ages, genders, races, or CKD stages. Furthermore, our study revealed that compared to non-LCD individuals, a higher proportion of LCD adherents have higher incomes. This may be due to the fact that individuals with higher incomes are relatively more likely to embrace new dietary concepts and have the energy to improve their lifestyle habits.

The long-term compliance to LCD may be lower compared to a normal diet. While individuals undergoing Very Low Calorie Diet (VLCD) or LCD treatments believe that this dietary pattern significantly benefits their conditions ^23^, providing them with a certain intrinsic motivation, the transition from unrestricted eating to LCD—requiring careful calculation for each meal — can negatively impact their emotional well-being, especially during dining out, as commercial meals are often high in carbohydrates, thus social behaviors may significantly limit adherence to LCD. Nielsen et al. conducted an LCD study involving 48 diabetic individuals, and found that after two years, approximately half of them had discontinued the LCD ^24^. The long-term maintenance of LCD adherence is influenced by multiple factors. Ahola et al. ^25^ researched the LCD compliance of 902 individuals with Type 1 Diabetes and discovered that lower BMI, glycemic variability, and diastolic blood pressure could affect long-term adherence. In our study, we found that younger age, Non-Hispanic Black, and Non-Hispanic White individuals were more likely to choose LCD. Specifically, younger individuals demonstrated a greater acceptance of new dietary concepts and innovative recipes ^26^. ^29^. In Hispanic diets, a major source of carbohydrates is rice, while in the diets of Non-Hispanic Blacks and Whites, rice accounts for only a small proportion of carbohydrate intake ^27^. Consequently, reducing carbohydrate intake is more readily accepted by Non-Hispanic Black and Non-Hispanic White individuals. Therefore, we need to pay more attention to dietary issues and conduct targeted education for CKM individuals. When advocating for LCD, it is crucial to consider individuals’ food preferences, adjust carbohydrate structures according to personal circumstances, enhance intrinsic motivation, and reduce external pressures, thereby optimizing the likelihood of long-term success.

LCD has been shown to be closely associated with various disease prognoses, but its impact on prognosis presents inconsistent and contradictory results across different studies. There is evidence suggesting that LCD has a protective effect against all-cause mortality; however, other studies have revealed potential negative effects, such as possible nutritional imbalances and muscle mass decline, adversely affecting patient health. Research in populations with Type 2 Diabetes (T2DM), CVD, and CKD has suggested that the use of LCD can reduce the risk of mortality. Hu et al. ^28^ included 10,101 T2DM individuals to explore the association between LCD patterns and mortality, finding that for every 10% increase in LCD adherence, the overall mortality rate decreased by 12%, demonstrating a significant negative correlation. Ren et al. ^29^ found that LCD intake in CKD individuals could lower all-cause mortality risk. This aligns with our research results, where we found that LCD was associated with a reduced risk of all-cause mortality in CKM individuals. However, a 15-year follow-up study on elderly individuals found no association between overall LCD scores and all-cause or cause-specific mortality risk ^30^. Seidelmann et al. studied 15,428 adults in four American communities, finding a “U” shaped relationship between carbohydrate intake and all-cause mortality, stating that both excessively high and low carbohydrate intakes led to increased mortality ^31^. Mazid et al. ^32^ based on the NHANES cohort study, reported a detrimental association between long-term LCD and overall mortality. These discrepancies compared to our findings may be because CKM individuals require LCD support for improved blood sugar control, optimized lipid metabolism, reduced inflammation, and better vascular and renal health ^33^, whereas the general population’ s long-term LCD might lead to a deficiency in essential nutrients, thereby increasing the risk of adverse outcomes.

Stratified analysis revealed that this relationship was not influenced by age, gender, race, presence of hypertension, or underlying CVD, and notably, results across different stages of CKM were consistent as well in our study. However, our study also found that the relationship between LCD and all-cause mortality in CKM patients exhibits heterogeneity depending on the presence of diabetes. Numerous studies have confirmed that LCD can effectively improve glycemic control, optimize lipid profiles, and alleviate metabolic syndrome-related indicators in patients with T2DM, thereby reducing their all-cause mortality rate ^34^. However, when examining individuals with CKM and concurrent diabetes, we found that such benefits were not significant. This finding suggests that not all types of CKM patients benefit from LCD, particularly in the presence of diabetes, where a more cautious approach may be necessary when applying LCD. This heterogeneity may be attributed to the failure of most studies to differentiate between types of carbohydrates when investigating LCD. In fact, within low-carbohydrate diets, different types and qualities of carbohydrates have varying effects on individual health ^31, 35^. A well-structured LCD should not contain excessive refined grains. When implementing LCD in patients with diabetes, special attention should be paid to avoid such dietary structure in the macronutrient profile ^36^. There is evidence indicating that patients exhibit a greater acute glycemic response following the fine milling of grains ^37^. A large multinational prospective cohort study conducted in 21 countries confirmed an association between increased consumption of white rice and heightened diabetes risk ^38^. Various national dietary guidelines ^39^ and diabetes management recommendations ^40^ encourage the consumption of whole grains and recommend substituting whole grains for refined grains. A follow-up study conducted by Sun et al. over 15 years found that replacing 50 grams per day of refined grains with whole grains could reduce the overall and CVD mortality risk by 4-5% in patients^41^. Therefore, although our results indicate that the LCD is beneficial in reducing all-cause mortality risk in patients with CKM, the increased risk of all-cause mortality associated with a low-quality LCD structure might act as a confounding factor, potentially obscuring the LCD’s true effects and leading to non-significant statistical results. Therefore, whether there exists further internal heterogeneity among CKM individuals with diabetes, as well as the most appropriate macro-nutrient combinations for CKM patients, requires further research for confirmation.

The LCD pattern may reduce the risk of all-cause mortality in CKM patients through various mechanisms. Firstly, by reducing carbohydrate intake, LCD can significantly improve multiple CVD risk factors, such as lowering triglycerides (TGs), optimizing low-density lipoprotein cholesterol (LDL-C) particle distribution^42^, and decreasing the individual’s weight, waist circumference, BMI, and body fat^43^. Secondly, there is a complex association between CKM and inflammation; elevated systemic inflammation indices can lead to insulin resistance and disrupted lipid metabolism, thus increasing cardiovascular risk factors ^44^. Numerous studies have indicated that LCD can also improve insulin resistance, reduce blood glucose fluctuation levels, leading to improvements in metabolic syndrome and cardiovascular risk factors in obese patients. Finally, some studies have shown that CKM may result from shared genetic risks that alter mitochondrial metabolism across multiple organs ^45^. LCD typically increases the production of ketone bodies (such as β-hydroxybutyrate), which can serve as an alternative energy source to glucose. Research indicates that β-hydroxybutyrate can have a beneficial effect on lifespan by inhibiting the activity of the inflammatory complex NLRP3, thereby reducing the increases of age-related pro-inflammatory cytokines ^46^. Moreover, ketone bodies possess antioxidant properties, reducing the production of reactive oxygen species (ROS) and improving adverse events associated with heart failure through mechanisms such as enhancing ATP production, normalizing mitochondrial function, and reducing oxidative stress ^47^. However, more extensive research is needed to explore the specific influencing factors and mechanisms of LCD therapy in CKM individuals.

Despite the efforts made in exploring related themes in this study, certain limitations are inevitably present. Firstly, we only selected dietary survey results from a single day for analysis, which does not reflect the dynamic dietary intake trajectories of individuals. Secondly, there may have been insufficient control of potential confounders such as the percentage of energy obtained from carbohydrates and variations in dietary quality, which may have influenced the research results to some degree. Additionally, as NHANES did not comprehensively cover evaluation indices directly related to CKM diagnosis, this may introduce certain biases in the prevalence estimates. Lastly, although we calibrated the model and performed sensitivity analyses, it remains challenging to balance the biases that may arise from all factors. Therefore, future research urgently requires more prospective randomized controlled trials to further validate the effectiveness of LCD concerning CKM outcome measures and to clearly define the optimal LCD patterns and suitable intervention time points.

In summary, this study confirms the relationship between LCD and the risk of all-cause mortality in CKM individuals, providing evidence to enhance understanding of LCD and CKM, as well as to guide individualized patient education and dietary management strategies for CKM. Future research should further validate the effectiveness of LCD concerning CKM outcome measures, aiming to provide more precise and effective guidance for dietary management in CKM individuals.

## Funding

The Science and technology fund of Chengdu Medical College (CYZYB22-02).

The research fund of Sichuan Medical and Health Care Promotion institute (KY2022QN0309).

Sichuan Provincial Medical Association Youth Innovation Project (Q23021).

Health Commission of Sichuan Province Medical Science and Technology Program (24QNMP100).

Sichuan Provincial Geriatric Clinical Medicine Research Center (24LHLNYX1-48).

## Author Contributions

Conception and design of the study: Zhuoxing Li, Xin Ma, Mao Xiao, Xue Zhang, Yanyi Deng, Yun Sun, Xiang Xiao;

Acquisition and analysis of data: Zhuoxing Li, Xin Ma;

Drafting the manuscript or figures: Zhuoxing Li, Xin Ma, Mao Xiao, Xue Zhang, Yanyi Deng, Yun Sun, Xiang Xiao.

## Data Availability

Some or all datasets generated or analyzed during the current study are not publicly available. However, they can be obtained from the corresponding author upon reasonable request.

## Acknowledgments

The authors express their gratitude to all participants of this study for their significant contributions.

## Ethical Approval

The study protocol adhered to the ethical standards set forth in the 1964 Declaration of Helsinki and its subsequent amendments. Approval was obtained from the National Committee for Ethical Review of Health Statistics Research, and all participants provided signed informed consent.

## Declarations

The authors state that there are no known financial conflicts of interest or personal relationships that could have influenced the research presented in this paper.

---

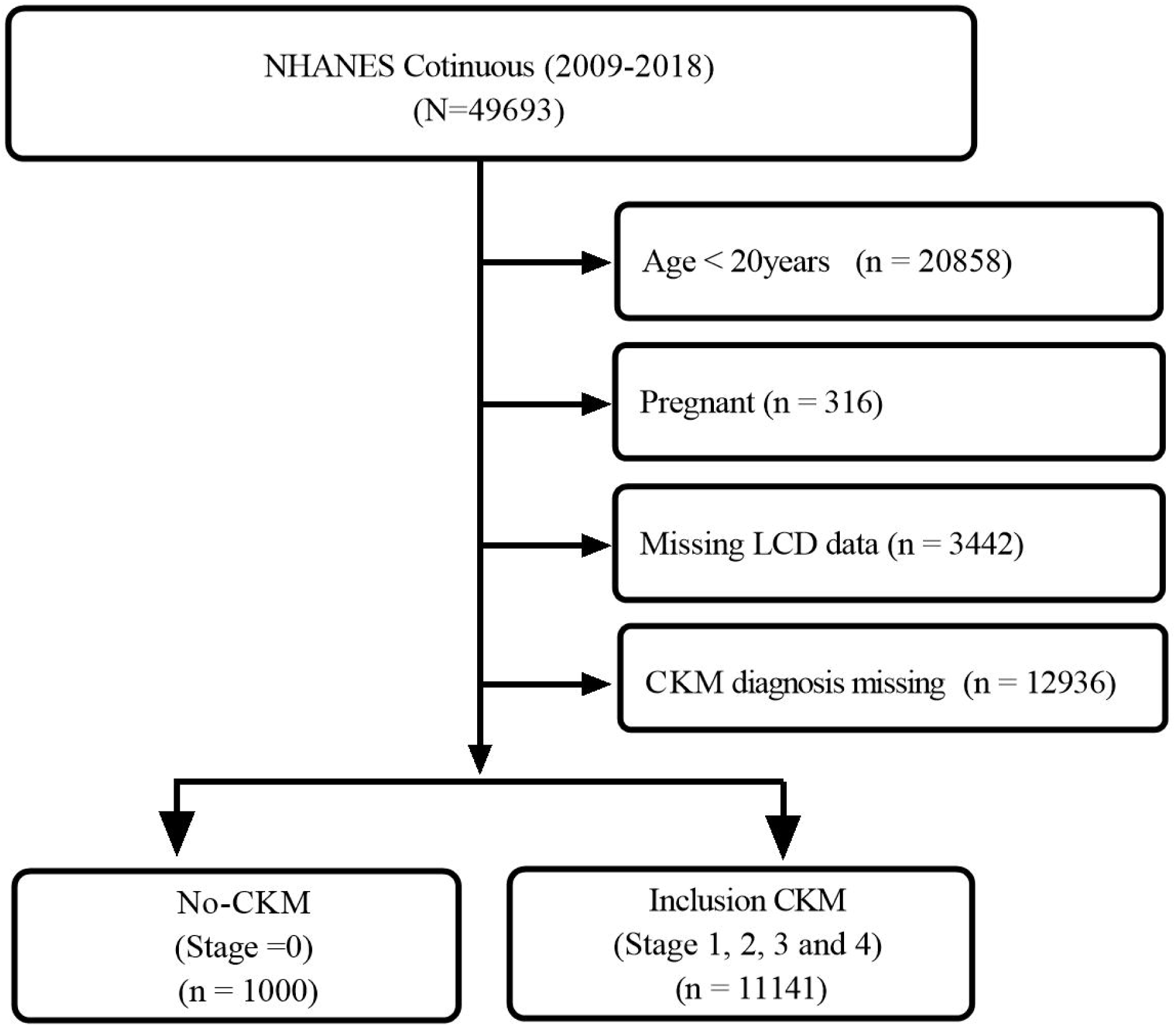

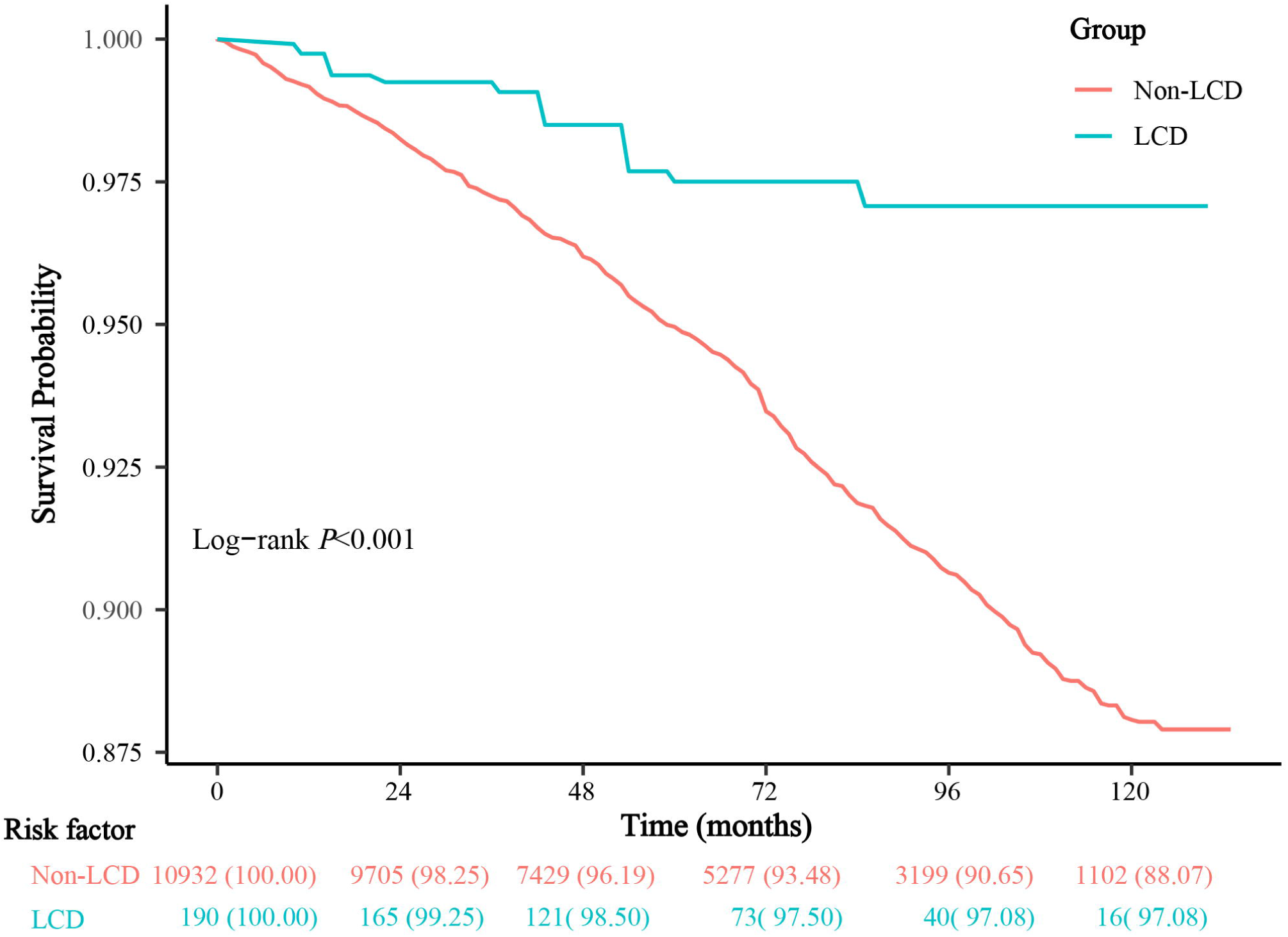

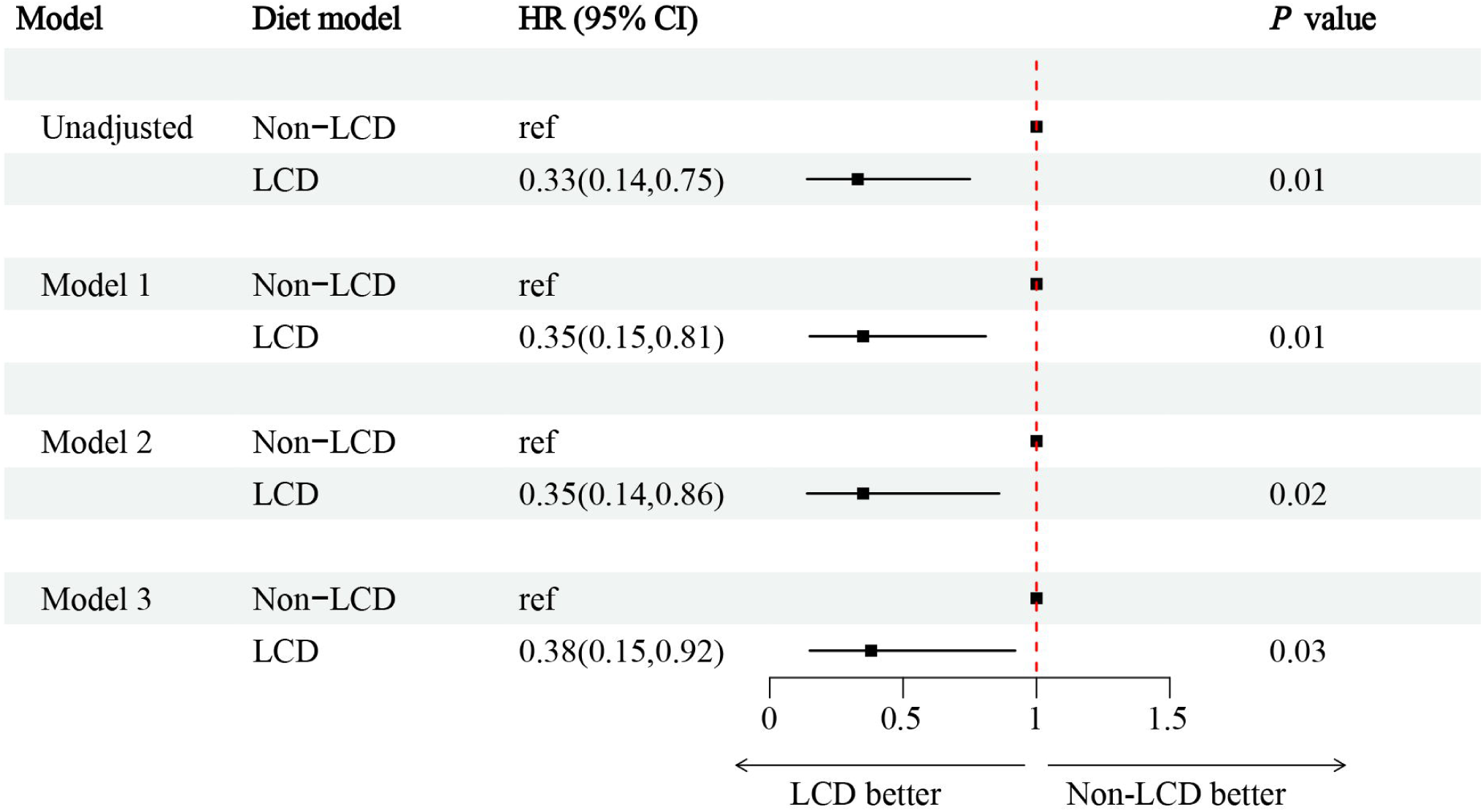

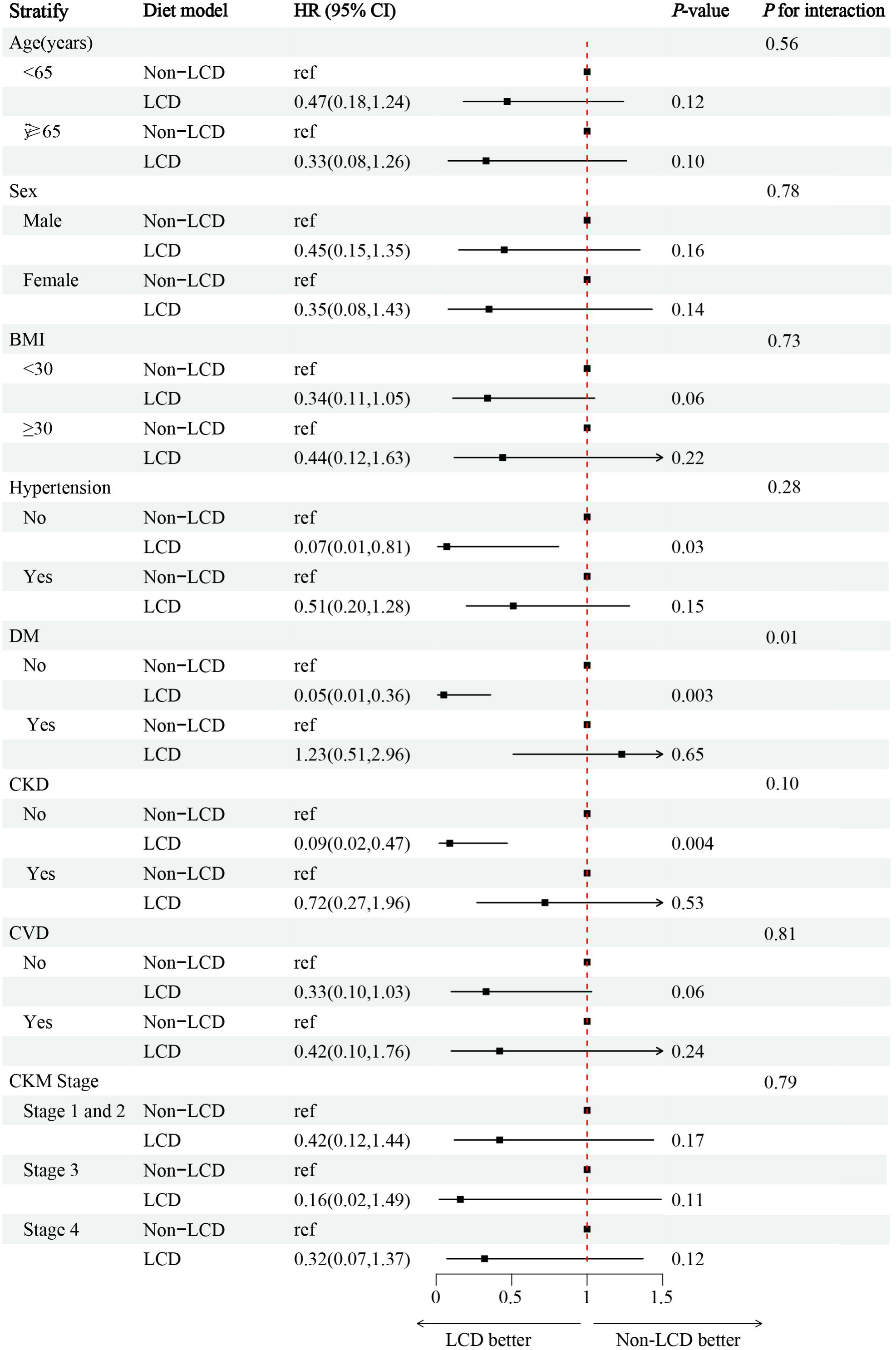

