## Supplementary material for "The Potential of Dietary Strategies: The Impact of Low-Carbohydrate Diet on Cardiovascular-Kidney-Metabolic Syndrome": Table 1&Table 2

**Table 1. Baseline clinical features of enrolled individuals with CKM**

| Variable | Total  (N=11141) | Non-LCD  (N=10950) | LCD  (N=191) | *P*-value |
| --- | --- | --- | --- | --- |
| Age (years) | 50.04(0.30) | 50.06(0.30) | 49.30(1.38) | 0.58 |
| Age (years) (%) |  |  |  | 0.13 |
| <65 | 77.59(0.02) | 77.47(0.65) | 83.02(3.24) |  |
| ≥65 | 22.41(0.01) | 22.53(0.65) | 16.98(3.24) |  |
| Sex (%) |  |  |  | 0.38 |
| Female | 49.76(0.01) | 49.66(0.65) | 54.51(5.43) |  |
| Male | 50.24(0.02) | 50.34(0.65) | 45.49(5.43) |  |
| Ethnicity (%) |  |  |  | **0.16** |
| Mexican American | 8.80(0.01) | 8.91(0.88) | 3.04(1.12) |  |
| Non-Hispanic Black | 11.51(0.01) | 11.46(0.88) | 13.87(2.70) |  |
| Non-Hispanic White | 65.09(0.03) | 65.00(1.58) | 69.21(4.67) |  |
| Other Hispanic | 6.24(0.01) | 6.22(0.59) | 7.01(2.31) |  |
| Other Race - Including Multi-Racial | 8.37(0.01) | 8.40(0.54) | 6.88(2.78) |  |
| BMI (kg/m²) | 29.98(0.12) | 29.98(0.12) | 29.92(0.61) | 0.92 |
| BMI (kg/m²) (%) |  |  |  | 0.79 |
| ≤30 | 57.72(0.02) | 57.75(0.75) | 56.35(5.23) |  |
| ＞30 | 42.28(0.01) | 42.25(0.75) | 43.65(5.23) |  |
| Hypertension (%) |  |  |  | 0.53 |
| No | 55.72(0.02) | 55.79(0.81) | 52.30(5.67) |  |
| Yes | 44.28(0.01) | 44.21(0.81) | 47.70(5.67) |  |
| Alcohol user (%) |  |  |  | **0.07** |
| No | 11.23(0.01) | 11.33(0.52) | 6.40(2.20) |  |
| Yes | 88.77(0.03) | 88.67(0.52) | 93.60(2.20) |  |
| Smoke (%) |  |  |  | 0.27 |
| No | 53.28(0.01) | 53.40(0.83) | 47.42(5.30) |  |
| Yes | 46.72(0.02) | 46.60(0.83) | 52.58(5.30) |  |
| Poverty (10,000 dollars) (%) |  |  |  | **0.05** |
| ≤1 | 21.54(0.01) | 21.53(0.91) | 21.72(4.86) |  |
| 1.1-3 | 34.89(0.01) | 35.13(0.85) | 23.19(4.42) |  |
| >3 | 43.57(0.02) | 43.33(1.26) | 55.08(5.85) |  |
| Educational level (%) |  |  |  | 0.09 |
| College or above | 59.75(0.02) | 59.58(1.18) | 69.85(4.93) |  |
| High school or equivalent | 34.70(0.01) | 34.90(1.06) | 26.00(4.62) |  |
| Less than high school | 5.49(0.00) | 5.52(0.39) | 4.15(1.70) |  |
| Anemia (%) |  |  |  | 0.50 |
| No | 92.83(0.03) | 92.81(0.35) | 94.20(1.84) |  |
| Yes | 7.17(0.00) | 7.19(0.35) | 5.80(1.84) |  |
| Hyperlipidemia (%) |  |  |  | 0.43 |
| No | 26.44(0.01) | 26.52(0.68) | 22.40(4.70) |  |
| Yes | 73.56(0.02) | 73.48(0.68) | 77.60(4.70) |  |
| HbA1c (%) | 5.72(0.01) | 5.72(0.01) | 5.59(0.08) | 0.07 |
| DM (%) |  |  |  | 0.23 |
| No | 81.13(0.02) | 81.05(0.60) | 84.72(2.93) |  |
| Yes | 18.87(0.01) | 18.95(0.60) | 15.28(2.93) |  |
| ACR (mg/g) | 38.66(2.76) | 38.66( 2.78) | 38.83(10.09) | 0.99 |
| ACR (mg/g) (%) |  |  |  | 0.82 |
| <30 | 89.08(0.03) | 89.10(0.42) | 87.80(3.19) |  |
| (30-300) | 9.02(0.00) | 9.00(0.36) | 9.99(3.06) |  |
| ≥300 | 1.90(0.00) | 1.89(0.16) | 2.21(0.82) |  |
| eGFR (ml/min/1.73m²) | 91.29(0.41) | 91.30(0.41) | 91.00(1.74) | 0.86 |
| e-GFR (ml/min/1.73m²) (%) |  |  |  | 0.21 |
| ≥60 | 89.28(0.03) | 89.22(0.44) | 91.96(1.93) |  |
| <60 | 10.72(0.00) | 10.78(0.44) | 8.04(1.93) |  |
| CKD (%) |  |  |  | 0.82 |
| No | 80.19(0.02) | 83.69(0.51) | 82.85(3.74) |  |
| Yes | 15.65(0.01) | 16.31(0.51) | 17.15(3.74) |  |
| CKM Syndrome (%) |  |  |  | 0.41 |
| Stage1 | 24.66(0.01) | 24.54(0.70) | 30.41(6.16) |  |
| Stage2 | 60.00(0.02) | 60.05(0.73) | 57.75(5.64) |  |
| Stage3 | 4.24(0.00) | 4.28(0.29) | 2.60(0.79) |  |
| Stage4 | 11.10(0.01) | 11.13(0.43) | 9.24(2.54) |  |
| CVD (%) |  |  |  | 0.58 |
| No | 89.22(0.02) | 89.19(0.43) | 90.76(2.54) |  |
| Yes | 10.78(0.01) | 10.81(0.43) | 9.24(2.54) |  |

CKM, Cardio-Kidney-Metabolic; LCD, Low-Carbohydrate Diet; BMI, Body Mass Index; ACR, albumin-to-creatinine ratio; DM, diabetes mellitus; e-GFR, estimated glomerular filtration rate; CKD, chronic kidney disease; CVD, Cardiovascular disease.

**Table 2. Prevalence of LCD in individuals with CKM**

| Variables | Non-LCD | LCD | *P* Value |
| --- | --- | --- | --- |
| Age (years) |  |  | 0.13 |
| <65 | 97.84(0.27) | 2.16(0.27) |  |
| ≥65 | 98.47(0.29) | 1.53(0.29) |  |
| Sex |  |  | 0.38 |
| Female | 97.79(0.32) | 2.21(0.32) |  |
| Male | 98.17(0.30) | 1.83(0.30) |  |
| Ethnicity |  |  | 0.16 |
| Mexican American | 99.30(0.249) | 0.70(0.249) |  |
| Non-Hispanic Black | 97.57(0.441) | 2.43(0.441) |  |
| Non-Hispanic White | 97.85(0.289) | 2.15(0.289) |  |
| Other Hispanic | 97.73(0.719) | 2.27(0.719) |  |
| Other Race - Including Multi-Racial | 98.34(0.695) | 1.66(0.695) |  |
| CKM Syndrome |  |  | 0.54 |
| Stage 0 | 98.39(0.65) | 1.61(0.65) |  |
| Stage 1 | 97.51(0.58) | 2.49(0.58) |  |
| Stage 2 | 98.06(0.27) | 1.94(0.27) |  |
| Stage 3 | 98.76(0.38) | 1.24(0.38) |  |
| Stage 4 | 98.32(0.47) | 1.68(0.47) |  |

LCD, Low-Carbohydrate Diet; CKM, Cardiovascular-kidney-Metabolic.

**Figure 1. Inclusion flowchart.**

**Figure 2. The relationship between LCD and the prognosis of individuals with CKM through Kaplan-Meier survival analysis.** LCD, Low-Carbohydrate Diet; CKM, Cardiovascular-kidney-Metabolic.

**Figure 3. Association between LCD and all-cause mortality in individuals with CKM. Model 1** adjusted for baseline age (‘<65’ years, ‘≥65’ years), gender (‘Female’, ‘Male’), ethnicity (‘Mexican American’, ‘Non-Hispanic Black’, ‘Non-Hispanic White’, ‘Other Hispanic’, ‘Other Race - Including Multi-Racial’), BMI (<30kg/m^2^, ≥30 kg/m^2^); **Model 2** adjusted for covariates in model 1 plus smoke (‘No’ or ‘Yes’), alcohol use (‘No’ or ‘Yes’), education (‘College or above’, ‘High school or equivalent’, ‘Less than high school’), poverty (‘0-1’, ‘1.1-3’, ‘＞3’) . **Model 3** adjusted for covariates in model 2 plus anemia (‘No’ or ‘Yes’), hyperlipidemia (‘No’ or ‘Yes’), hypertension (‘No’ or ‘Yes’), CKD (‘No’ or ‘Yes’), DM (‘No’ or ‘Yes’), CVD (‘No’ or ‘Yes’). LCD, Low-Carbohydrate Diet; CKM, Cardiovascular-kidney-Metabolic; HR, Hazard ratio; CI, Confidence interval; BMI, Body Mass Index; CVD, Cardiovascular disease; CKD, chronic kidney disease; DM, diabetes mellitus.

**Figure 4. Stratified analysis.** adjusted for baseline age (‘<65’ years, ‘≥65’ years), gender (‘Female’, ‘Male’), ethnicity (‘Mexican American’, ‘Non-Hispanic Black’, ‘Non-Hispanic White’, ‘Other Hispanic’, ‘Other Race - Including Multi-Racial’), BMI (<30kg/m^2^, ≥30 kg/m^2^), smoke (‘No’ or ‘Yes’), alcohol use (‘No’ or ‘Yes’), education (‘College or above’, ‘High school or equivalent’, ‘Less than high school’), poverty (‘0-1’, ‘1.1-3’, ‘＞3’). HR, Hazard ratio; CI, Confidence interval; BMI, Body Mass Index; CKD, chronic kidney disease; DM, diabetes mellitus; CVD, Cardiovascular disease; CKM, Cardiovascular-kidney-Metabolic.
