## Supplementary table 1-7 for "The Potential of Dietary Strategies: The Impact of Low-Carbohydrate Diet on Cardiovascular-Kidney-Metabolic Syndrome"

**Supplementary table 1. Definitions/criteria of some diagnoses**

| Variables | Definitions/criteria |
| --- | --- |
| Smoker | Smoking more than 100 cigarettes in previous and now. |
| Alcohol user ^[1]^ | ≥2 drinks per day for females, ≥3 drinks per day for males, or binge drinking ≥2 days per month.  Binge drinking (≥4 drinks on the same occasion for females, ≥5 drinks on the same occasion for males) on 5 or more days per month. |
| Hypertension ^[2]^ | (1) Self-reported hypertension diagnosis, (2) Use of anti-hypertensive medication, (3) Average systolic blood pressure (SBP) > 140 mmHg, (4) Average diastolic blood pressure (DBP) > 90 mmHg, meet any of the above conditions. |
| Anemia ^[3]^ | ≥120g/L for women (15 years of age and above), ≥130g/L for men (15 years of age and above). |
| Hyperlipidemia | (1) Triglyceridemia ≥ 150 mg/dl; (2) Hypercholesterolemia: a) total cholesterol ≥ 200 mg/dl, b) low-density lipoprotein ≥ 130 mg/dl), c). high-density lipoprotein (< 40 mg/dl, male; < 50 mg/dl, female), meet any of the above conditions; (3) Use of lipid-lowering drugs; meet any of the above conditions. |
| Chronic Kidney Disease (CKD) | Individuals with an albumin-to-creatinine ratio (ACR) higher than 30 mg/g and/or an estimated glomerular filtration rate (eGFR) lower than 60 mL/min/1.73 m^2^ were defined as CKD patients ^[4]^. |
| Cardiovascular Disease | Coronary heart disease; congestive heart failure; heart attack; stroke; angina. |
| Low-carbohydrate diet ^[5]^ | Very low-carbohydrate (<10% carbohydrates) or 20 to 50 g/d  Low-carbohydrate (<26% carbohydrates) or less than 130 g/d  Moderate-carbohydrate (26%-44%)  High-carbohydrate (45% or greater) |
| Cardio-Kidney-Metabolic (CKM) Syndrome ^[6]^ | We define CKM stage 0 as non-CKM, and CKM stages 1, 2, 3, and 4 as CKM.  **Stage 0: No CKM health risk factors**  Participants classified under CKM Stage 0 exhibited a normal body mass index (BMI) (less than 23 kg/m² for Asian individuals and less than 25 kg/m² for other racial and ethnic groups) and a normal waist circumference (less than 80 cm for Asian women and less than 90 cm for Asian men, and less than 88 cm for women and less than 102 cm for men from all other racial and ethnic categories, respectively). These individuals did not meet the criteria for any other stages.  **Stage 1: Excess and/or dysfunctional adiposity**  CKM Stage 1 encompassed individuals with an increased BMI (23 kg/m² or more for Asians and over 25 kg/m² for all other racial and ethnic groups), an increased waist circumference (80 cm or more for Asian women and 90 cm or more for Asian men, and 88 cm or more for women and 102 cm or more for men from other racial and ethnic categories, respectively), or prediabetes (glycated hemoglobin levels of 5.7% to less than 6.5% or fasting blood glucose levels of 100 mg/dL to less than 126 mg/dL).  **Stage 2: Metabolic risk factors and CKD**  According to Kidney Disease Improving Global Outcomes (KDIGO) criteria, as recommended by the AHA, CKM Stage 2 identified participants with metabolic risk factors or moderate-to-high-risk chronic kidney disease (CKD). These metabolic risk factors included elevated fasting serum triglycerides (135 mg/dL or more), hypertension, diabetes, or metabolic syndrome (presence of at least three of the following: increased waist circumference, low high-density lipoprotein cholesterol (HDL) levels [<40 mg/dL for men and <50 mg/dL for women], fasting serum triglycerides of 150 mg/dL or more, elevated blood pressure [systolic blood pressure of 130 mmHg or more, diastolic blood pressure of 80 mmHg or more, and/or use of blood pressure-lowering medications], or prediabetes). CKD stages were determined based on glomerular filtration rate (GFR) and urinary albumin-to-creatinine ratio.  **Stage 3: Subclinical CVD in CKM**  CKM Stage 3 was identified by the presence of very-high-risk KDIGO CKD stages or a high predicted 10-year cardiovascular disease (CVD) risk. The 10-year CVD risk was estimated using the AHA Predicting Risk of CVD EVENTs (PREVENT) equations. High risk was defined as a 10-year CVD risk of 20% or more (based on recommended thresholds). The PREVENT equations were developed and validated for adults aged 30 to 79 years, excluding adults younger than 30 years from risk estimation. However, to avoid underestimating CKD Stage 3, adults aged 80 years or more were not excluded from the 10-year CVD risk assessment. Instead, they were assigned an age of 79 years for conservative risk estimates. Furthermore, PREVENT was designed for variables within specific ranges: total cholesterol (130-320 mg/dL), HDL (20-100 mg/dL), systolic blood pressure (90-200 mmHg), and GFR (14-140 mL/min/1.73m²). Values for these variables outside these bounds were adjusted to the allowable upper or lower limits (e.g., a total cholesterol of 330 mg/dL was set to 320 mg/dL). Cardiac biomarkers and cardiovascular imaging were not used to identify subclinical CVD.  **Stage 4: Clinical CVD in CKM**  CKM Stage 4 was determined based on self-reported established cardiovascular diseases, including coronary heart disease, angina, heart attack, heart failure, and stroke. Atrial fibrillation and peripheral artery disease were excluded due to the unavailability of these data. |

**Supplementary-table 2. Logistic regression analysis to assess the factors influencing the choice of a LCD in patients with CKM**

| Variables | Univariate Logistic Regression | | Multivariate Logistic Regression | |
| --- | --- | --- | --- | --- |
|  | OR (95% CI) | *P* value | OR (95% CI) | *P* value |
| Age (years) |  |  |  |  |
| <65 | ref | ref | ref | ref |
| ≥65 | 0.70(0.45,1.11) | 0.13 | 0.57(0.36,0.91) | **0.02** |
| Gender |  |  |  |  |
| Male | ref | ref | ref | ref |
| Female | 0.82(0.53,1.28) | 0.38 | 0.78(0.49,1.24) | 0.29 |
| BMI (kg/m^2^) |  |  |  |  |
| <18.5 | ref | ref | ref | ref |
| ≥30 | 1.06(0.69,1.63) | 0.79 | 1.03(0.65,1.65) | 0.89 |
| Ethnicity |  |  |  |  |
| Mexican American | ref | ref | ref | ref |
| Non-Hispanic Black | 3.54(1.56,8.03) | **0.003** | 3.59(1.47,8.75) | **0.01** |
| Non-Hispanic White | 3.12(1.44,6.75) | **0.004** | 2.74(1.19,6.35) | **0.02** |
| Other Hispanic | 3.30(1.27,8.58) | **0.02** | 2.72(1.00,7.44) | **0.05** |
| Other Race - Including Multi-Racial | 2.40(0.77,7.50) | 0.13 | 2.06(0.62,6.83) | 0.23 |
| Alcohol user |  |  |  |  |
| No | ref | ref | ref | ref |
| Yes | 1.87(0.93,3.74) | 0.08 | 1.52(0.73,3.19) | 0.26 |
| Smoke |  |  |  |  |
| No | ref | ref | ref | ref |
| Yes | 1.27(0.83,1.96) | 0.27 | 1.30(0.84,2.01) | 0.24 |
| Poverty (10,000 dollars) |  |  |  |  |
| 0-1 | ref | ref | ref | ref |
| 1.1-3 | 0.65(0.35,1.23) | 0.18 | 0.69(0.35,1.39) | 0.30 |
| >3 | 1.26(0.70,2.26) | 0.43 | 1.21(0.61,2.40) | 0.58 |
| Educational level |  |  |  |  |
| College or above | ref | ref | ref | ref |
| High school or equivalent | 0.64(0.40,1.01) | 0.06 | 0.72(0.43,1.20) | 0.20 |
| Less than high school | 0.64(0.27,1.52) | 0.31 | 1.05(0.40,2.76) | 0.92 |
| Anemia |  |  |  |  |
| No | ref | ref | ref | ref |
| Yes | 0.79(0.40,1.57) | 0.50 | 0.90(0.43,1.91) | 0.78 |
| Hyperlipidemia |  |  |  |  |
| No | ref | ref | ref | ref |
| Yes | 1.25(0.71,2.19) | 0.43 | 1.25(0.71,2.22) | 0.43 |
| Hypertension |  |  |  |  |
| No | ref | ref | ref | ref |
| Yes | 1.15(0.74,1.80) | 0.53 | 1.17(0.72,1.90) | 0.52 |
| CKD |  |  |  |  |
| No | ref | ref | ref | ref |
| Yes | 1.06(0.64,1.76) | 0.82 | 1.38(0.78,2.44) | 0.26 |
| DM |  |  |  |  |
| No | ref | ref | ref | ref |
| Yes | 0.77(0.50,1.18) | 0.23 | 0.77(0.46,1.27) | 0.30 |
| CVD |  |  |  |  |
| No | ref | ref | ref | ref |
| Yes | 0.84(0.45,1.57) | 0.58 | 0.95(0.51,1.77) | 0.86 |

LCD, Low-Carbohydrate Diet ;CKM, Cardio-Kidney-Metabolic; OR, Odds ratio; CI, Confidence interval; BMI, Body Mass Index; CKD, chronic kidney disease; DM, diabetes mellitus; CVD, Cardiovascular disease.

**Supplementary -table 3. Cox-regression analysis of risk factors for all-cause mortality in individuals with CKM**

| Variables | Unadjusted | | Model 1 | | Model 2 | | Model 3 | |
| --- | --- | --- | --- | --- | --- | --- | --- | --- |
|  | HR (95% CI) | *P*-value | HR (95% CI) | *P* value | HR (95% CI) | *P* value | HR (95% CI) | *P* value |
| LCD |  |  |  |  |  |  |  |  |
| No | ref |  | ref | ref | ref | ref | ref | ref |
| Yes | 0.33(0.14,0.75) | 0.01 | 0.35(0.15,0.81) | 0.01 | 0.35(0.14,0.86) | 0.02 | 0.38(0.15,0.92) | 0.03 |
| Age (years) |  |  |  |  |  |  |  |  |
| <65 | ref | ref | ref |  | ref | ref | ref | ref |
| ≥65 | 7.37(5.95,9.13) | <0.0001 | 8.15(6.66,9.97) | <0.0001 | 7.55(6.04,9.43) | <0.0001 | 3.86(2.95,5.05) | <0.0001 |
| Sex |  |  |  |  |  |  |  |  |
| Female | ref | ref | ref |  | ref | ref | ref | ref |
| Male | 1.16(0.99,1.35) | 0.07 | 1.32(1.13,1.55) | <0.0001 | 1.33(1.13,1.57) | <0.0001 | 1.35(1.12,1.63) | 0.002 |
| Ethnicity |  |  |  |  |  |  |  |  |
| Mexican American | ref | ref | ref | ref | ref | ref | ref | ref |
| Non-Hispanic Black | 1.87(1.33,2.64) | <0.001 | 1.62(1.20,2.18) | 0.002 | 1.97(1.45,2.66) | <0.0001 | 1.34(0.98,1.83) | 0.06 |
| Non-Hispanic White | 2.23(1.61,3.09) | <0.0001 | 1.35(1.01,1.82) | 0.04 | 2.00(1.46,2.74) | <0.0001 | 1.81(1.34,2.45) | <0.001 |
| Other Hispanic | 0.83(0.57,1.22) | 0.35 | 0.76(0.54,1.07) | 0.11 | 0.83(0.59,1.17) | 0.30 | 0.83(0.56,1.23) | 0.34 |
| Other Race - Including Multi-Racial | 1.19(0.75,1.88) | 0.46 | 0.90(0.59,1.39) | 0.64 | 1.22(0.79,1.87) | 0.37 | 0.92(0.61,1.39) | 0.71 |
| BMI (kg/m2) |  |  |  |  |  |  |  |  |
| <30 | ref | ref | ref | ref | ref | ref | ref | ref |
| ≥30 | 0.95(0.79,1.14) | 0.57 | 1.05(0.86,1.27) | 0.64 | 1.05(0.87,1.26) | 0.63 | 0.85(0.70,1.05) | 0.13 |
| Alcohol use |  |  |  |  |  |  |  |  |
| No | ref | ref |  |  | ref | ref | ref | ref |
| Yes | 0.77(0.60,0.98) | 0.04 |  |  | 1.57(1.30,1.90) | <0.0001 | 0.97(0.75,1.25) | 0.81 |
| Smoke |  |  |  |  |  |  |  |  |
| No | ref | ref |  |  | ref | ref | ref | ref |
| Yes | 1.87(1.54,2.26) | <0.0001 |  |  | 1.54(1.33,1.77) | <0.001 | 1.43(1.18,1.75) | <0.001 |
| Poverty (10000 dollar) (%) |  |  |  |  |  |  |  |  |
| ≤1 | ref | ref |  |  | ref | ref | ref | ref |
| 1.1-3 | 1.29(0.96,1.71) | 0.09 |  |  | 0.92(0.70,1.21) | 0.54 | 0.94(0.71,1.23) | 0.64 |
| >3 | 0.61(0.47,0.80) | <0.0001 |  |  | 0.52(0.39,0.69) | <0.0001 | 0.54(0.41,0.72) | <0.0001 |
| Education (%) |  |  |  |  |  |  |  |  |
| High school or equivalent | ref | ref |  |  | ref | ref | ref | ref |
| Less than high school | 1.64(1.32,2.03) | <0.0001 |  |  | 1.29(1.02,1.62) | 0.03 | 1.16(0.92,1.46) | 0.22 |
| College or above | 2.59(1.86,3.60) | <0.0001 |  |  | 1.82(1.24,2.66) | 0.002 | 1.44(0.98,2.11) | 0.06 |
| Anemia |  |  |  |  |  |  |  |  |
| No | ref | ref |  |  |  |  | ref | ref |
| Yes | 4.19(3.53,4.97) | <0.0001 |  |  |  |  | 2.22(1.77,2.79) | <0.0001 |
| Hyperlipidemia |  |  |  |  |  |  |  |  |
| No | ref | ref |  |  |  |  | ref | ref |
| Yes | 1.64(1.30,2.06) | <0.0001 |  |  |  |  | 1.13(0.86,1.48) | 0.39 |
| Hypertension |  |  |  |  |  |  |  |  |
| No | ref | ref |  |  |  |  | ref | ref |
| Yes | 3.95(3.27,4.77) | <0.0001 |  |  |  |  | 1.62(1.25,2.10) | <0.001 |
| DM |  |  |  |  |  |  |  |  |
| No | ref | ref |  |  |  |  | ref |  |
| Yes | 2.93(2.44,3.51) | <0.0001 |  |  |  |  | 1.22(0.99,1.51) | 0.06 |
| CKD |  |  |  |  |  |  |  |  |
| No | ref | ref |  |  |  |  | ref | ref |
| Yes | 6.38(5.30,7.68) | <0.0001 |  |  |  |  | 2.35(1.91,2.89) | <0.0001 |
| CVD |  |  |  |  |  |  |  |  |
| No | ref | ref |  |  |  |  | ref | ref |
| Yes | 5.43(4.48,6.58) | <0.0001 |  |  |  |  | 1.62(1.28,2.06) | <0.0001 |

**Model 1** adjusted for baseline age (‘<65’ years, ‘≥65’ years), gender (‘Female’, ‘Male’), race (‘Mexican American’, ‘Non-Hispanic Black’, ‘Non-Hispanic White’, ‘Other Hispanic’, ‘Other Race - Including Multi-Racial’), BMI (<30kg/m^2^, ≥30 kg/m^2^); **Model 2** adjusted for covariates in model 1 plus smoke (‘Yes’ or ‘No’), alcohol use (‘Yes’ or ‘No’), education (‘College or above’, ‘High school or equivalent’, ‘Less than high school’), poverty (‘0-1’, ‘1.1-3’, ‘＞3’) . **Model 3** adjusted for covariates in model 2 plus anemia (‘Yes’ or ‘No’), hyperlipidemia (‘Yes’ or ‘No’), hypertension (‘Yes’ or ‘No’), CKD (‘Yes’ or ‘No’), DM (‘Yes’ or ‘No’), CVD (‘Yes’ or ‘No’). LCD, Low-Carbohydrate Diet; CKM, Cardiovascular-kidney-Metabolic; HR, Hazard ratio; CI, Confidence interval; BMI, Body Mass Index; CVD, Cardiovascular disease; CKD, chronic kidney disease; DM, diabetes mellitus.

**Supplementary-table 4. The relationship between LCD and all-cause mortality in patients with CKM**

| Variables | Unadjusted |  | Model 1 |  | Model 2 |  | Model 3 |  |
| --- | --- | --- | --- | --- | --- | --- | --- | --- |
|  | HR (95%CI) | *P* value | HR (95%CI) | *P* value | HR (95%CI) | *P* value | HR (95%CI) | *P* value |
| Non-LCD | ref |  | ref |  | ref |  | ref |  |
| LCD | 0.33(0.14,0.75) | 0.01 | 0.35(0.15,0.81) | 0.01 | 0.35(0.14,0.86) | 0.02 | 0.38(0.15,0.92) | 0.03 |

**Model 1** adjusted for baseline age (‘<65’ years, ‘≥65’ years), gender (‘Female’, ‘Male’), race (‘Mexican American’, ‘Non-Hispanic Black’, ‘Non-Hispanic White’, ‘Other Hispanic’, ‘Other Race - Including Multi-Racial’), BMI (<30kg/m^2^, ≥30 kg/m^2^); **Model 2** adjusted for covariates in model 1 plus smoke (‘Yes’ or ‘No’), alcohol use (‘Yes’ or ‘No’), education (‘College or above’, ‘High school or equivalent’, ‘Less than high school’), poverty (‘0-1’, ‘1.1-3’, ‘＞3’) . **Model 3** adjusted for covariates in model 2 plus anemia (‘Yes’ or ‘No’), hyperlipidemia (‘Yes’ or ‘No’), hypertension (‘Yes’ or ‘No’), CKD (‘Yes’ or ‘No’), DM (‘Yes’ or ‘No’), CVD (‘Yes’ or ‘No’). LCD, Low-Carbohydrate Diet; CKM, Cardiovascular-kidney-Metabolic; HR, Hazard ratio; CI, Confidence interval; BMI, Body Mass Index; CKD, chronic kidney disease; DM, diabetes mellitus; CVD, Cardiovascular disease.

**Supplementary-table 5. Stratified analysis of LCD and the risk of all-cause mortality in individuals with CKM**

| Variable | HR（95% CI） | | | *P* for interaction |
| --- | --- | --- | --- | --- |
|  | Non-LCD | LCD | *P* for trend |  |
| Age (years) |  |  |  | 0.56 |
| <65 | ref | 0.47(0.18,1.24) | 0.13 |  |
| ≥65 | ref | 0.33(0.08,1.26) | 0.10 |  |
| Gender |  |  |  | 0.78 |
| Male | ref | 0.45(0.15,1.35) | 0.16 |  |
| Female | ref | 0.35(0.08,1.43) | 0.14 |  |
| BMI (kg/m2) |  |  |  | 0.73 |
| ≤30 | ref | 0.34(0.11,1.05) | 0.06 |  |
| ＞30 | ref | 0.44(0.12,1.63) | 0.22 |  |
| Hypertension |  |  |  | 0.28 |
| No | ref | 0.07(0.01,0.81) | **0.03** |  |
| Yes | ref | 0.51(0.20,1.28) | 0.15 |  |
| DM |  |  |  | **0.01** |
| No | ref | 0.05(0.01,0.36) | **0.003** |  |
| Yes | ref | 1.23(0.51,2.96) | 0.65 |  |
| CKD |  |  |  | 0.10 |
| No | ref | 0.09(0.02,0.47) | **0.004** |  |
| Yes | ref | 0.72(0.27,1.96) | 0.53 |  |
| CVD |  |  |  | 0.81 |
| No | ref | 0.33(0.10,1.03) | **0.06** |  |
| Yes | ref | 0.42(0.10,1.76) | 0.24 |  |
| CKM Syndrome |  |  |  | 0.79 |
| Stage 1-2 | ref | 0.42(0.12,1.44) | 0.17 |  |
| Stage 3 | ref | 0.16(0.02,1.49) | 0.11 |  |
| Stage 4 | ref | 0.32(0.07,1.37) | 0.12 |  |

adjusted for baseline age (‘<65’ years, ‘≥65’ years), gender (‘Female’, ‘Male’), race (‘Mexican American’, ‘Non-Hispanic Black’, ‘Non-Hispanic White’, ‘Other Hispanic’, ‘Other Race - Including Multi-Racial’), BMI (<30kg/m^2^, ≥30 kg/m^2^), smoke (‘Yes’ or ‘No’), alcohol use (‘Yes’ or ‘No’), education (‘College or above’, ‘High school or equivalent’, ‘Less than high school’), poverty (‘0-1’, ‘1.1-3’, ‘＞3’). HR, Hazard ratio; CI, Confidence interval; BMI, Body Mass Index; CKD, chronic kidney disease; DM, diabetes mellitus; CVD, Cardiovascular disease; CKM, Cardiovascular-kidney-Metabolic.

**Supplementary-table 6. The relationship between LCD and all-cause mortality in patients with CKM after deleting missing values**

| Variables | Unadjusted |  | Model 1 |  | Model 2 |  | Model 3 |  |
| --- | --- | --- | --- | --- | --- | --- | --- | --- |
|  | HR (95%CI) | *P* value | HR (95%CI) | *P* value | HR (95%CI) | *P* value | HR (95%CI) | *P* value |
| Non-LCD | ref |  | ref |  | ref |  | ref |  |
| LCD | 0.33(0.14,0.75) | 0.01 | 0.36(0.15,0.83) | 0.02 | 0.30(0.11,0.82) | 0.02 | 0.33(0.13,0.85) | 0.02 |

**Model 1*^a^*** adjusted for baseline age (‘<65’ years, ‘≥65’ years), gender (‘Female’, ‘Male’), race (‘Mexican American’, ‘Non-Hispanic Black’, ‘Non-Hispanic White’, ‘Other Hispanic’, ‘Other Race - Including Multi-Racial’), BMI (<30kg/m^2^, ≥30 kg/m^2^); **Model 2*^b^*** adjusted for covariates in model 1 plus smoke (‘Yes’ or ‘No’), alcohol use (‘Yes’ or ‘No’), education (‘College or above’, ‘High school or equivalent’, ‘Less than high school’), poverty (‘0-1’, ‘1.1-3’, ‘＞3’) . **Model 3*^c^*** adjusted for covariates in model 2 plus anemia (‘Yes’ or ‘No’), hyperlipidemia (‘Yes’ or ‘No’), hypertension (‘Yes’ or ‘No’)，DM (‘Yes’ or ‘No’), CKM, Cardiovascular-kidney-Metabolic; HR, Hazard ratio; CI, Confidence interval; BMI, Body Mass Index; CKD, chronic kidney disease; DM, diabetes mellitus, CVD, Cardiovascular disease.

**Supplementary-table 7. Exploring the relationship between LCD and all-cause mortality in individuals with CKM after excluding gastrointestinal cancer, COPD, and depression**

| Variables | Unadjusted |  | Model 1 |  | Model 2 |  | Model 3 |  |
| --- | --- | --- | --- | --- | --- | --- | --- | --- |
|  | HR (95%CI) | *P* value | HR (95%CI) | *P* value | HR (95%CI) | *P* value | HR (95%CI) | *P* value |
| Non-LCD | ref |  | ref |  | ref |  | ref |  |
| LCD | 0.34(0.15,0.78) | 0.01 | 0.36(0.15,0.83) | 0.02 | 0.35(0.14,0.87) | 0.02 | 0.38(0.15,0.91) | 0.03 |

**Model 1*^a^*** adjusted for baseline age (‘<65’ years, ‘≥65’ years), gender (‘Female’, ‘Male’), race (‘Mexican American’, ‘Non-Hispanic Black’, ‘Non-Hispanic White’, ‘Other Hispanic’, ‘Other Race - Including Multi-Racial’), BMI (<30kg/m^2^, ≥30 kg/m^2^); **Model 2*^b^*** adjusted for covariates in model 1 plus smoke (‘Yes’ or ‘No’), alcohol use (‘Yes’ or ‘No’), education (‘College or above’, ‘High school or equivalent’, ‘Less than high school’), poverty (‘0-1’, ‘1.1-3’, ‘＞3’) . **Model 3*^c^*** adjusted for covariates in model 2 plus anemia (‘Yes’ or ‘No’), hyperlipidemia (‘Yes’ or ‘No’), hypertension (‘Yes’ or ‘No’)，DM (‘Yes’ or ‘No’), CKM, Cardiovascular-kidney-Metabolic; COPD, Chronic Obstructive Pulmonary Disease**;** HR, Hazard ratio; CI, Confidence interval; BMI, Body Mass Index; CKD, chronic kidney disease; DM, diabetes mellitus, CVD, Cardiovascular disease.

[5]	OH R, GILANI B, UPPALURI K R. Low-Carbohydrate Diet [M]. StatPearls. Treasure Island (FL) ineligible companies. Disclosure: Brian Gilani declares no relevant financial relationships with ineligible companies. Disclosure: Kalyan Uppaluri declares no relevant financial relationships with ineligible companies.; StatPearls Publishing

Copyright © 2024, StatPearls Publishing LLC. 2024.

[6]	Aggarwal R, Ostrominski JW, Vaduganathan M. Prevalence of Cardiovascular-Kidney-Metabolic Syndrome Stages in US Adults, 2011-2020. JAMA. 2024 Jun 4;331(21):1858-1860.
